# Mechanism of bisphosphonate-related osteonecrosis of the jaw (BRONJ) revealed by targeted removal of legacy bisphosphonate from jawbone using equilibrium competing inert hydroxymethylene diphosphonate

**DOI:** 10.1101/2021.12.22.21268006

**Authors:** Hiroko Okawa, Takeru Kondo, Akishige Hokugo, Philip Cherian, Jesus J. Campagna, Nicholas Lentini, Shuting Sun, Eric C. Sung, Samantha Chiang, Yi-Ling Lin, Frank H. Ebetino, Varghese John, Charles E. McKenna, Ichiro Nishimura

**Affiliations:** Weintraub Center for Reconstructive Biotechnology, Division of Advanced Prosthodontics, UCLA School of Dentistry; Los Angeles, CA 90095; Department of Molecular & Regenerative Prosthodontics, Tohoku University Graduate School of Dentistry; Sendai, Japan 980-8575; Division of Plastic & Reconstructive Surgery, David Geffen School of Medicine at UCLA; Los Angeles, CA 90095; BioVinc, LLC; Pasadena, CA 91107; Department of Neurology, David Geffen School of Medicine at UCLA; Los Angeles, CA 90095; Department of Chemistry, University of Southern California; Los Angeles, CA 90089; Section of Oral Biology, UCLA School of Dentistry; Los Angeles, CA 90095; Section of Oral & Maxillofacial Pathology, UCLA School of Dentistry; Los Angeles, CA 90095

## Abstract

Bisphosphonate-related osteonecrosis of the jaw (BRONJ) presents as a morbid jawbone lesion in patients exposed to a nitrogen-containing bisphosphonate (N-BP). Although it is rare, BRONJ has caused apprehension among patients and healthcare providers and decreased acceptance of this anti-resorptive drug class to treat osteoporosis and metastatic osteolysis. We report here a novel method to elucidate the pathological mechanism of BRONJ by the selective removal of legacy N-BP from the jawbone using an intra-oral application of hydroxymethylene diphosphonate (HMDP) formulated in deformable nanoscale vesicles (DNV). After maxillary tooth extraction, zoledronate-treated mice developed delayed gingival wound closure, delayed tooth extraction socket healing and increased jawbone osteonecrosis consistent with human BRONJ lesion. Single cell RNA sequencing of mouse gingival cells revealed oral barrier immune dysregulation and unresolved pro-inflammatory reaction. HMDP-DNV topical applications to nascent mouse BRONJ lesions resulted in accelerated gingival wound closure and bone socket healing as well as attenuation of osteonecrosis development. The gingival single cell RNA sequencing demonstrated resolution of chronic inflammation by increased anti-inflammatory signature gene expression of lymphocytes and myeloid-derived suppressor cells. This study suggests that BRONJ pathology was predominantly induced by the oral N-BP and demonstrates the potential of HMDP-DNV as an effective BRONJ therapy.

**Brief Summary:** The targeted removal of legacy bisphosphonate from the jawbone by competitive equilibrium therapy elucidated the pathological mechanism of aberrant oral barrier immunity and bisphosphonate-related osteonecrosis of the jaw (BRONJ)

## INTRODUCTION

Nitrogen-containing bisphosphonates (N-BPs) are prototypical anti-resorptive agents (1–3) initially marketed to prevent bone fractures and to treat osteopenia or osteoporosis (4, 5). N-BPs have been widely prescribed for postmenopausal women who have a bone mineral density T score of −2.5 or less, a history of spine or hip fracture, or a Fracture Risk Assessment Tool score indicating increased fracture risk. N-BP treatment with an increased dose and frequency was also given to patients with multiple myeloma (6) or metastatic cancers to bone marrow (7, 8) to address tumor-induced osteolysis, hypercalcemia and bone pain. N-BPs are well tolerated, exhibit few side-effects, and have established clinical benefits (5, 9).

In the 2000’s, cases of osteonecrosis in the jawbone (ONJ) emerged among a minority of patients with a history of N-BP treatment, usually occurring at a dental infection site (10) or after dentoalveolar surgery such as a tooth extraction (11, 12). Originally designated “Bisphosphonate-related ONJ” (BRONJ), this syndrome has also proven to be associated with non-bisphosphonate antiresorptive agents such as denosumab, a humanized anti-RANKL monoclonal antibody and anti-angiogenesis drugs. As a result, the American Association for Oral & Maxillofacial Surgeons (AAOMS) has proposed the term “Medication-related ONJ” (MRONJ) reflecting the association of ONJ with a multiplicity of antiresorptive agents (13).

Incidents of MRONJ reported to the United States Food and Drug Administration (FDA)’s Adverse Event Reporting System (FAERS) peaked from the first quarter of 2010 to the first quarter of 2014 with approximately 30,000 cases during this period, among which BRONJ represented the major fraction (14). The FARES database may underreport the incidents because of variations in provider’s perceptions of how severe an event needs to be to warrant submission of the event report (15). We present an alternative MRONJ prevalence estimation of approximately 36,000 new cases per year in the US (Table 1), which is still within the FDA’s orphan disease definition of less than 200,000 cases annually (16).

**Table 1.** Estimated MRONJ case numbers in the US based on MRONJ prevalence for the major underlying diseases*

| **Underlying Diseases** | **New Cases in the US (Year)** | **MRONJ Prevalence** | **Estimated MRONJ Cases (Year)\*** |
| --- | --- | --- | --- |
| Multiple Myeloma | 34,920** | 5.16% (89) | 1,802 |
| Breast Cancer | 330,840*** | 2.09% (89) | 6,915 |
| Prostate Cancer | 248,530** | 3.80% (89) | 9,444 |
| Osteoporosis | 1,800,000**** | 0.01% (90) | 18,000 |
| **Estimated Annual Incidents of MRONJ** |  |  | **36,161** |

\*Based on an assumption that all these patients were treated by anti-resorptive medications.
\*\*American Cancer Society. *Cancer Facts & Figures 2021*. Atlanta, Ga: American Cancer Society; 2021
\*\*\*Invasive breast cancer and ductal carcinoma: American Cancer Society. How Common Is Breast Cancer? Jan. 2020. Available at: https://www.cancer.org/cancer/breast-cancer/about/how-common-is-breast-cancer.html.
\*\*\*\*Cases in 2020 estimated from 2013 data: National Osteoporosis Foundation at: https://www.nof.org/news/54-million-americans-affected-by-osteoporosis-and-low-bone-mass/

Although the reported case prevalence is small, long-lasting severe clinical symptoms of MRONJ in some patients (Fig. S1) have created apprehension among patients and healthcare providers (17, 18). The NIH Workshop on “Pathway to Prevention (P2P) for Osteoporotic Fracture” in 2018 highlighted the acute need of research to mitigate serious adverse events such as MRONJ and atypical femur fracture to prevent the increasing threat of osteoporotic fractures (19). The P2P workshop particularly highlighted the markedly decreased acceptance of anti-resorptive medications by osteoporosis patients, despite the significant benefit of these drugs for this indication (20). The recent decline in N-BP prescription, reflecting diminished patient acceptance of these drugs, has been linked to a statistical rise in bone-related complications (21). Due to the long-lasting half-life of N-BPs once chemisorbed to bone, the discontinuation of N-BPs and switching to other anti-resorptive medications may still pose a risk of developing BRONJ (22). It is therefore urgent to reduce the BRONJ risk associated with these otherwise effective medications for pathological osteolysis.

The mechanism of BRONJ is not yet fully understood (23). Characterization of BRONJ patients suggested that the autoimmune diseases or systemic immunosuppression might contribute to the pathogenesis. Clinical case studies proposed an increased risk of BRONJ in patients with rheumatoid arthritis (24), autoimmune hepatitis (25), and Sjogren syndrome (26, 27). However, a study of larger BRONJ cohort did not show a clear relationship (28). Autoimmune diseases are often treated with immunosuppressive agents, which might affect the susceptibility of BRONJ (29, 30). N-BP-treated rodent models combined with experimental rheumatoid arthritis (31) or concurrent dexamethasone treatment (32) developed exacerbated BRONJ lesion supporting the susceptibility of compromised systemic immunity.

A prospective study of human subjects diagnosed with multiple myeloma demonstrated the reduced neutrophil function and chemotaxis activity after the first intravenous (IV) infusion of N-BP (33). The reduced function and chemotaxis of neutrophils were reported in BRONJ patients as compared to healthy subjects or osteoradionecrosis patients (34). BRONJ patients were also reported to exhibit the deficiency of peripheral blood γδT cells (35). Repeated N-BP treatments may contribute to the deficiency in immune function, although the exact role of systemic immunosuppression has not been clearly linked to the initiation of BRONJ pathogenesis (36).

Typical disease symptoms include gingival dehiscence and exposure of necrotic jawbone, which persists over months to years (13, 37, 38). The necrotic jawbone often becomes infected, resulting in pain, erythema, and purulent drainage (39). BRONJ exclusively occurs in the jawbone, which uniquely associated with the gingival oral barrier tissue with one of the most active barrier immunities (40). The presence of N-BP on the jawbone is shared by all BRONJ patients (41) and it is conceivable that jawbone N-BP may uniquely interact with oral barrier immunity. The major challenge has been the difficulty in separating the systemic and oral effects of N-BP therapy, which has severly hindered the elucidation of the mechanisms of BRONJ.

N-BP chemisorption to bone results from its high affinity to hydroxyapatite (HAp), the mineral component of bone (42). Recent investigations in our laboratories revealed that pre-adsorbed N-BPs can be displaced from HAp by exposure to a second N-BP/BP in aqueous buffer both *in vitro* and *in vivo* (43, 44). This led us to envision a new experimental model based on the selective removal of pharmacologically active legacy N-BP locally from jawbone by intra-oral application of a second BP with low pharmacological potency (lpBP). Herein, we describe a liposome-based deformable nanoscale vesicle (DNV) formulation for topical trans-oral mucosa delivery of an lpBP to the underlying jawbone. Here we report its effect in attenuating N-BP-induced oral lesions using a murine BRONJ model and the elucidation of the mechanism involved in this pathology.

## RESULTS

### Hydroxymethylene diphosphonate (HMDP: hydroxymethylene bisphosphonate) was selected as the lpBP

N-BPs such as zoledronate (ZOL) act on osteoclasts by inhibiting farnesyl diphosphate synthase (FPPS), thus interfering with the mevalonate pathway and protein prenylation (45). The nitrogen-containing side chain of N-BPs, an essential pharmacophore in all potent N-BP anti-resorptive drugs, is absent in hydroxymethylene diphosphonate (HMDP) which however retains α-OH group together with two phosphonate groups, conferring high affinity to HAp (Fig. 1A). Thus, we postulated that HMDP in solution might be able to decrease the amount of legacy N-BP at the bone surface by a competitive displacement mechanism as we reported (43, 44). ZOL IV injection to mice increased femur trabecular bone volume over the saline vehicle solution injection control, whereas HMDP IV injection with the same dose did not alter the bone architecture (Fig. 1B and 1C). The lack of pharmacological effect of HMDP *in vivo* was consistent with our previous study using a standard *in vitro* prenylation assay (44) and confirmed HMDP to be an lpBP.

**Fig. 1.**
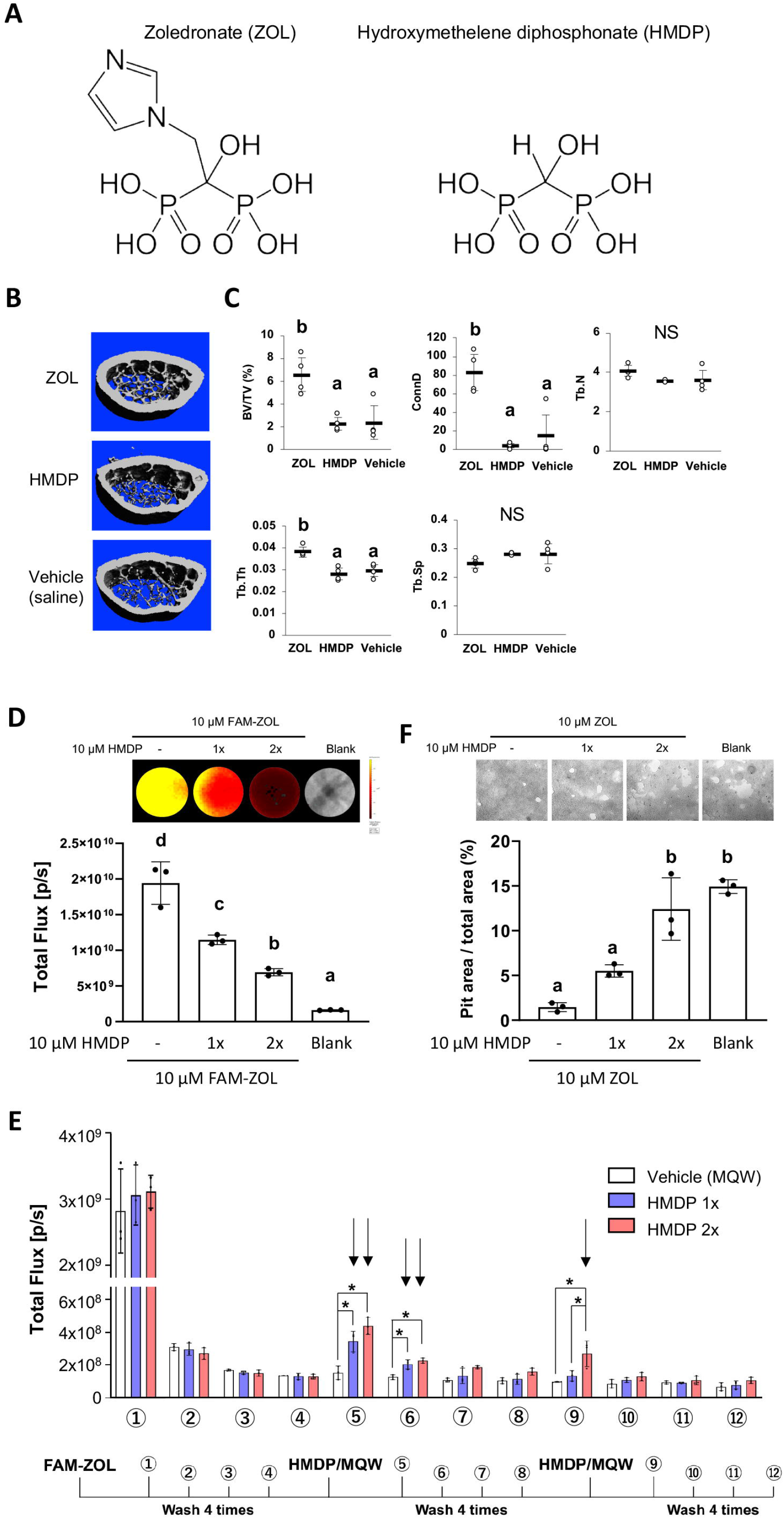
Competitive equilibrium-based dissociation of N-BP by low potency BP (lpBP). **(A)** Chemical structures (shown as the tetraacids) of N-BP, zoledronate (ZOL) and hydroxymethylene diphosphonate (HMDP). **(B)** ZOL but not HMDP affects mouse femur trabecular bone architecture. Mice received a bolus intravenous injection of ZOL (40 nmol), HMDP (40 nmol) or vehicle saline (0.9% NaCl) solutions. Femurs (n=4 per group) were harvested 3 weeks after the IV injection and subjected to Micro-CT imaging. **(C)** HMDP did not affect the femur trabecular bone micro-architecture, whereas ZOL increased bone volume over total volume (BV/TV), connectivity density (ConnD) and trabecular thickness (Tb.Th). Trabecular number (Tb.N) and trabecular separation (Tb.Sp) measurements were not affected by ZOL. **(D)** *In vitro* demonstration of competitive displacement of legacy N-BP. Synthetic apatite-coated wells pre-incubated with 10 µM FAM-ZOL were washed with MilliQ-treated pure water (MQW), then treated with 10 µM HMDP once (1x) or twice (2x) (n=3 per group). The FAM fluorescent signal measurement indicated significant reduction of the FAM-ZOL amount on the synthetic apatite. **(E)** Fluorescent signal measurement of the wash solutions from the *in vitro* experiment in (**D**) demonstrated removal of FAM-ZOL by the HMDP treatments (arrows). **(F)** *In vitro* osteoclastic pit formation assay. Synthetic apatite coated wells were pre-incubated with 10 µM ZOL followed by 10 µM HMDP treatment 1x or 2x (n=3 per group). RAW274.1 cells (2.5×10^4^ cells per well) were then inoculated to each well in culture medium supplemented by mouse recombinant receptor activator of nuclear kappa-B ligand (RANKL). The areas of resorption pits generated by osteoclasts derived from RAW 264.7 cells were measured after 6 days of incubation and cell removal. Twice repeated HMDP treatments restored normal *in vitro* resorption pit formation.

Next, we tested the ability of HMDP to displace the pre-adsorbed N-BP from bone mineral surface. Synthetic apatite (carbonate apatite)-coated culture wells were pre-treated with fluorescently tagged ZOL (FAM-ZOL: 10 µM). After thorough washing to remove non-chemisorbed FAM-ZOL, the wells were challenged by application of HMDP (10 µM in Milli-Q treated pure water: MQW). FAM-ZOL on the synthetic apatite was reduced by half (Fig. 1D), and the wash solution contained a signal of dissociated FAM-ZOL (Fig. 1E). After the second HMDP application, the FAM-ZOL on the synthetic apatite was further reduced (Fig. 1D). This experiment confirmed that a chemisorbed N-BP could be displaced by repeated applications of HMDP (Fig. 1D, 1E).

We further examined the effect of competitive removal of ZOL on osteoclastic bone resorption *in vitro*. ZOL chemisorbed on synthetic apatite inhibited osteoclastic bone resorption *in vitro* as expected, which was measured by the decreased resorption pit area created by RAW 264.7-derived osteoclasts (Fig. 1F). A single application of HMDP increased the pit area, albeit at no statistical significance. When HMDP was applied twice, the resorption pit size significantly increased and reached the level of ZOL-untreated blank wells (Fig. 1F). Although the FAM-ZOL experiment suggested that ZOL would not completely be removed after two applications of HMDP, osteoclastic activity was restored to a nearly normal level as ZOL-untreated control group (Fig. 1F). We postulate that there may be a threshold bioavailable concentration of N-BP causing osteoclast abnormality. Therefore, our goal need not to be complete removal of legacy N-BP from the jawbone, but rather to decrease the local N-BP concentration below a BRONJ-triggering threshold.

### Preparation of the HMDP-deformable nano-scale vesicle (DNV) formulation

In our previous proof-of-concept experiment, HMDP directly injected into mouse palatal gingiva in ZOL-pretreated mice prior to the maxillary first molar extraction was shown to prevent the development of BRONJ (44). BRONJ lesions exhibit ulcerative gingival tissue exposing necrotic jawbone (Fig. S1). Because injection into pliable oral tissue will be challenging in some cases, we designed a formulation of HMDP enabling the compound to penetrate through the oral mucosa epithelial layer, making topical delivery to the jawbone possible. Liposomes have often been used as a drug carrier for controlled delivery to enhance drug concentrations in targeted tissues and to achieve therapeutic effects using minimum drug doses (46). Deformable nano-scale vesicles (DNV) comprise a modified liposome drug carrier (Fig. 2A) synthesized by a controlled microfluidics system, which due to nanovesicle deformability allows penetration through the keratinized epithelial layer of skin (47).

**Fig. 2.**
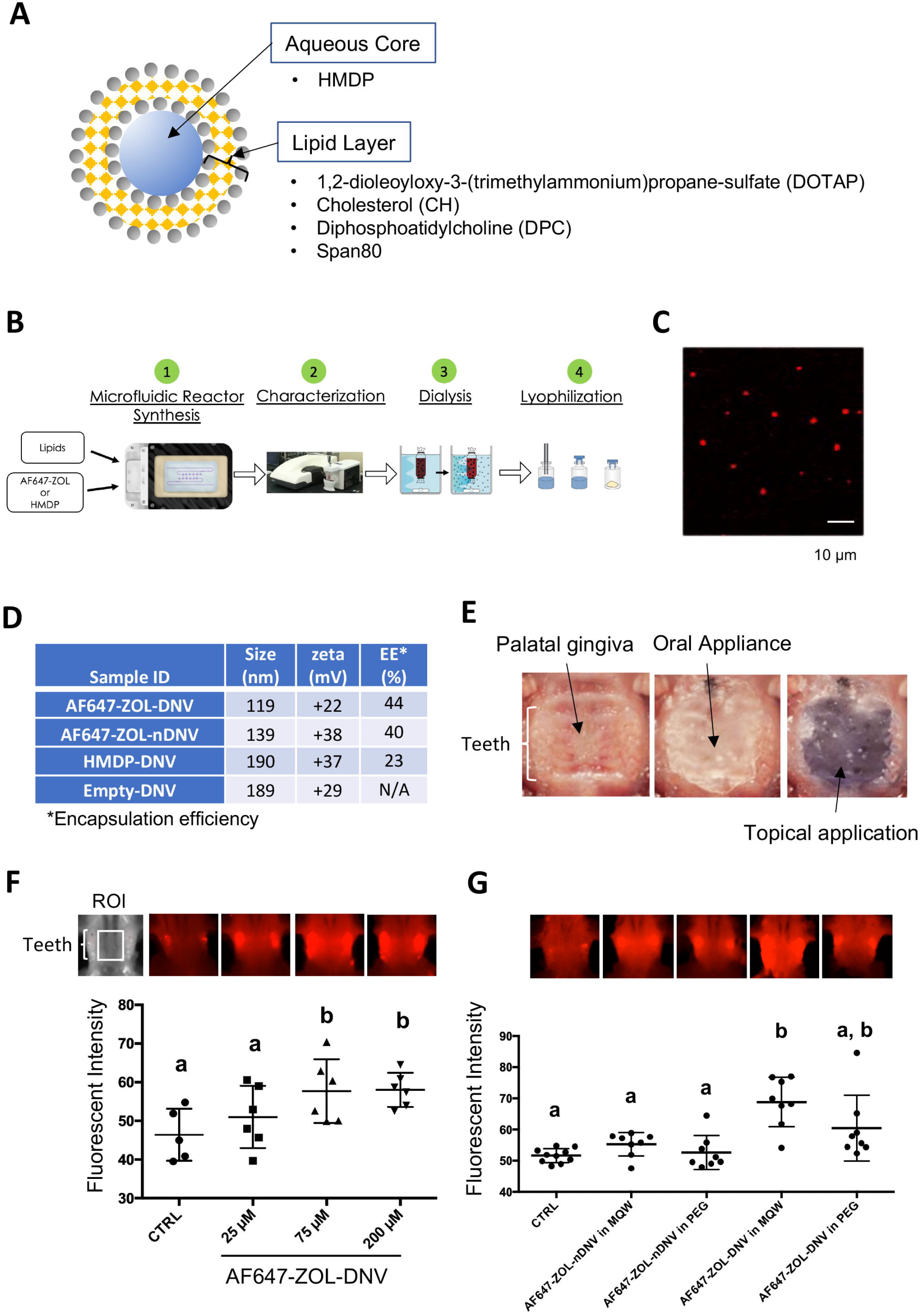
Manufacturing and trans-oral mucosal penetration evaluation of HMDP-DNV and AF647-ZOL-DNV. **(A)** Diagram of DNV, a liposome derivative. **(B)** Flow diagram of micro-fluidics based DNV synthesis. **(C)** Confocal laser scanning microscopy of AF647-ZOL-DNV. Approximately 100-200 nm DNV particles exhibited an AF647 signal. **(D)** Characterization of DNV formulations. **(E)** Protocol for intra-oral topical application to mouse palatal tissue. The reconstituted DNV solution in MQW (3 µl) was topically applied to the palatal gingiva between maxillary molar teeth and covered by a custom-made oral appliance fabricated with auto-polymerizing dental resin. After 1 hour, the oral appliance was removed. **(F)** After topical application of AF647-ZOL-DNV, mouse maxillary bones were harvested and AF647 fluorescence was measured. The AF647 fluorescent signal from the maxillary bone region of interest (ROI) increased with up to 75 µM AF7647-ZOL in DNV applied and then reached a plateau. **(G)** AF647-ZOL-DNV and AF647-ZOL in non-deformable formulation (AF647-ZOL-nDNV) were reconstituted in either MQW or 20% polyethylene glycol (PEG). AF647-ZOL-DNV in MQW most efficiently delivered the drug to the maxillary bone through trans-oral mucosal route.

HMDP-DNV contains HMDP in the aqueous core and lipid layers formed by 1,2-dioleoyloxy-3-(trimethylammonium)propane-sulfate (DOTAP), diphosphatidylcholine (DPPC), cholesterol (CH) and Span80 (15%v/v) (Fig. 2A). Span80 is a surfactant that generates DNV deformability. HMDP-DNV was manufactured through a synchronized stringent pipeline for microfluidic synthesis, followed by dialysis and freeze-drying (Fig. 2B).

DNV containing a far-red fluorophore (Alexa Fluor 647)-labeled ZOL (AF647-ZOL) (48) (BioVinc LLC, Pasadena, CA) was manufactured (Fig. 2C) to examine trans-oral mucosa drug delivery *in vivo*. We also synthesized “non-deformable” nanoscale vesicle (nDNV) without Span80 for use as a control.

The BP containing DNV sample was evaluated by particle size and surface zeta potential (Fig. 1D), which are reported to play important roles in liposome drug delivery behavior (49, 50). The optimal size for this purpose has been shown to be between 100 to 200 nm (47). After passing through the gingival and oral mucosa epithelial layers, DNV is expected to deliver HMDP to the bone mineral surface, which is negatively charged in isotonic solution (51). Chemisorption of N-BPs including ZOL further decreases the zeta potential of the functionalized bone mineral surface (52). Therefore, cationic DNV with a surface potential between +20 to +40 mV was selected to target N-BP-chemisorbed jawbone.

The trans-epithelial drug delivery of mouse palatal gingiva was designed to apply HMDP-DNV for 1 h. During application, the mouse palate was covered by a custom-made oral appliance using dental resin to protect from licking and accidental swallowing (Fig. 2E). In the initial study, lyophilized AF647-ZOL-DNV was dissolved in MQW at different concentrations (25 µM, 75µM and 200 µM) and 3 µl of the AF647-ZOL-DNV solution was applied to the mouse palatal gingival tissue. Forty-eight hours later, euthanized mouse skulls including the palatal alveolar bone were examined for fluorescent signal (Fig. 2F). AF647 fluorescent signal intensity at the maxillary bone increased with up to an applied concentration of 75 µM of AF647-ZOL and then reached a plateau.

We then prepared AF647-ZOL-DNV and AF647-ZOL-nDNV formulations, which were reconstituted in MQW or 20% polyethylene glycol (PEG). The AF647-ZOL-DNV (75 µM) reconstituted in MQW revealed the highest fluorescent signal, indicating successful AF647-ZOL delivery to the jawbone *in vivo* (Fig. 2G). This study also demonstrated that AF647-ZOL-nDNV (75 µM) was less effective in trans-oral mucosa delivery of AF647-ZOL to the jawbone.

### Development of a new mouse model: HMDP-DNV applications prior to dentoalveolar procedures in ZOL-treated mice

Female mice aged 8 to 10 weeks old were treated by a bolus ZOL intravenous (IV) injection (500 µg/Kg) from the retro-orbital venous plexus. Control mice received a vehicle solution IV injection. One week after the ZOL or vehicle injection, the maxillary left first molar was extracted (Fig. 3A). Extraction wound healing of the control group was uneventful, and the open gingival wound was mostly closed 1 week after the tooth extraction. By contrast, the tooth extraction wound of the ZOL group was delayed, which was evidenced by a gingival open wound until 4 weeks (Fig. 3B).

**Fig. 3.**
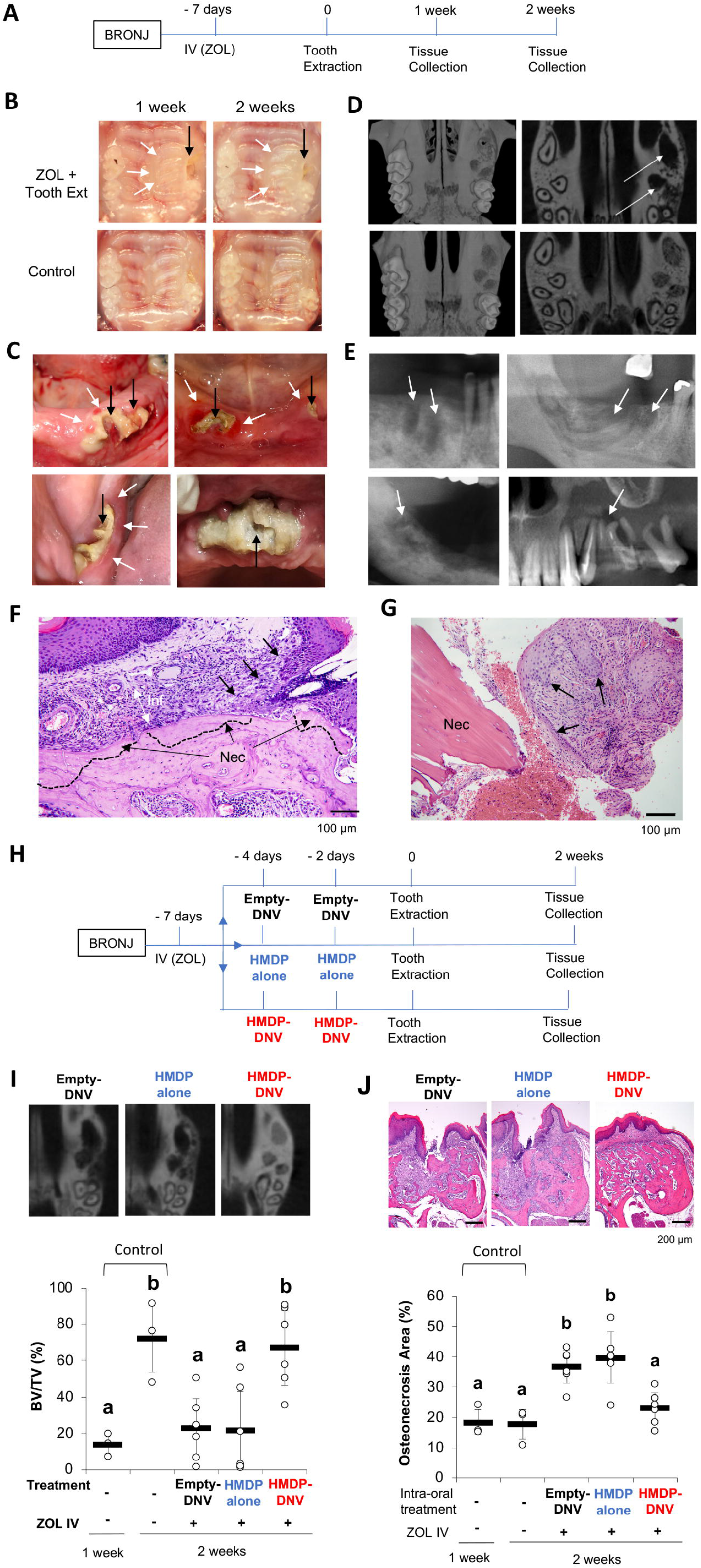
Disease phenotypes of mouse and human BRONJ lesion. **(A)** Experimental protocol for inducing a BRONJ lesion in mice. Mice received a bolus IV injection of ZOL (40 nmol/500 µg/Kg) or vehicle saline solution were subjected to maxillary left first molar extraction. **(B)** Intra-oral photographs depicted that mouse pretreated with ZOL IV injection exhibited delayed wound healing with sustained open wound (black arrows) and gingival swelling (white arrows). **(C)** Human BRONJ lesions with open tooth extraction wound (black arrows) and gingival swelling (white arrows). **(D)** Micro-CT images of mouse maxilla depicted that delayed bone regeneration in the mesial, buccal and palatal root extraction sockets (white arrows) in ZOL-pretreated mice, a sign of BRONJ symptoms. **(E)** Radiographic demonstration of unhealed tooth extraction of human BRONJ. **(F)** Histological evaluation of mouse BRONJ lesion with osteonecrosis (dotted line; Nec), gingival inflammation (Inf) and epithelial hyperplasia reaching to the necrotic bone (black arrows). **(G)** A biopsy specimen of human BRONJ lesion with osteonecrosis (Nec) and gingival epithelial hyperplasia (black arrows). **(H)** A time course experimental diagram of HMDP-DNV application. All mice received ZOL IV injection. Empty-DNV, HMDP in MQW (1.5 pmol/100 µM) or HMDP-DNV in MQW (1.5 pmol/100 µM) was topically applied to the palatal gingiva prior to the maxillary left first molar extraction. **(I)** Micro-CT evaluation. Two topical applications of HMDP-DNV prior to the tooth extraction significantly increased tooth extraction socket bone regeneration compared to Empty-DNV and HMDP alone. **(J)** Histological evaluation. Two topical applications of HMDP-DNV prior to the tooth extraction reduced the development of osteonecrosis compared to the treatment of Empty-DNV and HMDP alone, which remained to exhibit BRONJ phenotype.

The clinical definition of BRONJ is an unhealed oral wound with the exposed jawbone or the development of fistula reaching to the jawbone surface (Fig. 3C). AAOMS defined MRONJ including BRONJ as the nonhealing oral wound for 8 weeks in human patients (13) to differentiate the normal oral wound healing such as tooth extraction, which would be healed within this period. The tooth extraction wound in mice heals much faster in 3 weeks (36, 53). However, we found that the conventional mouse chaw pellets often delayed the tooth extraction wound healing due to food impaction in the extraction socket (44). Refining the mouse tooth extraction model by feeding soft gel diet after tooth extraction for 1 week, we found that the tooth extraction wound was clinically closed as early as 1 week (44). Using this refined mouse tooth extraction model, the BRONJ is assessed as open oral wound for 1 week of tooth extraction or later. The gross clinical observation is accomplished by using standardized oral photographs. The prevalence of open oral wound is expressed as the percent of animals with unhealed wound in each group at a given healing time.

After tooth extraction, the bony socket undergoes a sequential wound healing resulting in bone regeneration. Micro-CT is a well-established radiographic method suitable for small animal models. We have established a quantitative method to measure the bone volume in the mouse tooth extraction socket using Micro-CT images (Fig. S2). BRONJ delayed the bone regeneration process and often represented as empty tooth extraction socket in ZOL pretreated mice (Fig. 3D). A recent article reported the diagnostic value of radiographs for BRONJ (54), which exhibited radiographic features with unhealed empty extraction sockets and inflammatory lesion (Fig. 3E).

The development of osteonecrosis is the hallmark of BRONJ histopathology. In the mouse model, the necropsy of harvested maxillary tissues was conducted. The tooth extraction site of ZOL-injected mice exhibited a large area of osteonecrosis defined by a cluster of empty osteocytic lacunae. The gingival connective tissue adjacent to the necrotic bone showed dense inflammatory cell infiltration associated with epithelial hyperplasia (Fig. 3F). The bone biopsy specimens obtained from human BRONJ patients demonstrated the empty osteocytic lacunae as the definitive sign of osteonecrosis and associated with epithelial hyperplasia (Fig. 3G), reported as pseudoepitheliomatous hyperplasia (55).

Using the mouse BRONJ model, the effect of topical application of HMDP-DNV was examined to determine whether it would modulate the BRONJ symptoms. In this study, Empty-DNV, HMDP in MQW (1.5 pmol/100 µM) or HMDP-DNV in MQW (1.5 pmol/100 µM) were applied topically to the palatal tissue of mice pretreated with ZOL IV injection prior to the maxillary left first molar extraction (Fig. 3H). One time application of Empty-DNV, HMDP and HMDP-DNV did not affect the delayed tooth extraction wound healing in Micro-CT imaging and necropsy histological analyses (Fig. S3). Therefore, we increased the HMDP dose by twice topical applications prior to the tooth extraction. After 2x topical applications, the HMDP-DNV treated group showed increased bone regeneration at the equivalent level of the no ZOL-pretreated control group, whereas applications of Empty-DNV and HMDP alone had no significant effect (Fig. 3I).

Histological examination of the ‘HMDP-DNV 2x’-treated group showed normal extraction wound healing. The ‘Empty-DNV 2x’ and ‘HMDP alone 2x’-treated groups revealed extensive alveolar bone osteonecrosis, characteristic of a BRONJ lesion (Fig. 3J). The area of osteonecrosis in the maxillary alveolar bone in the ‘HMDP-DNV 2x’ application group was significantly prevented (Fig. 3J). Dentoalveolar surgeries such as tooth extraction have been reported as a risk factor to induced ONJ in patients with a history of N-BP therapy (13). The results suggest that the twice repeated applications of HMDP-DNV prior to dentoalveolar surgery (tooth extraction) prevented the development of BRONJ.

### Targeted removal of ZOL by HMDP-DNV treatment from nascent BRONJ lesions accelerated disease resolution in mice

BRONJ lesion in mice was defined as unhealed wound 1 week after tooth extraction in ZOL-injected mice, showing delayed gingival wound closure and exposed alveolar bone (Fig. 3B). Mouse maxillary tissues were harvested 1, 2 and 4 weeks after tooth extraction in the untreated BRONJ group (Fig. 4A). Twice repeated HMDP-DNV topical treatments were administered to the BRONJ lesion after 1 week of tooth extraction (Fig. 4A). Intra-oral examination revealed that the gingival wound was clinically closed in all mice that received the topical HMDP-DNV treatment, whereas the BRONJ lesion with an open gingival wound was observed in untreated mice (Fig. 4B). The area of open wound (Fig. 4C) and the area of gingival swelling (Fig. 4D) were significantly reduced by HMDP-DNV treatment.

**Fig. 4.**
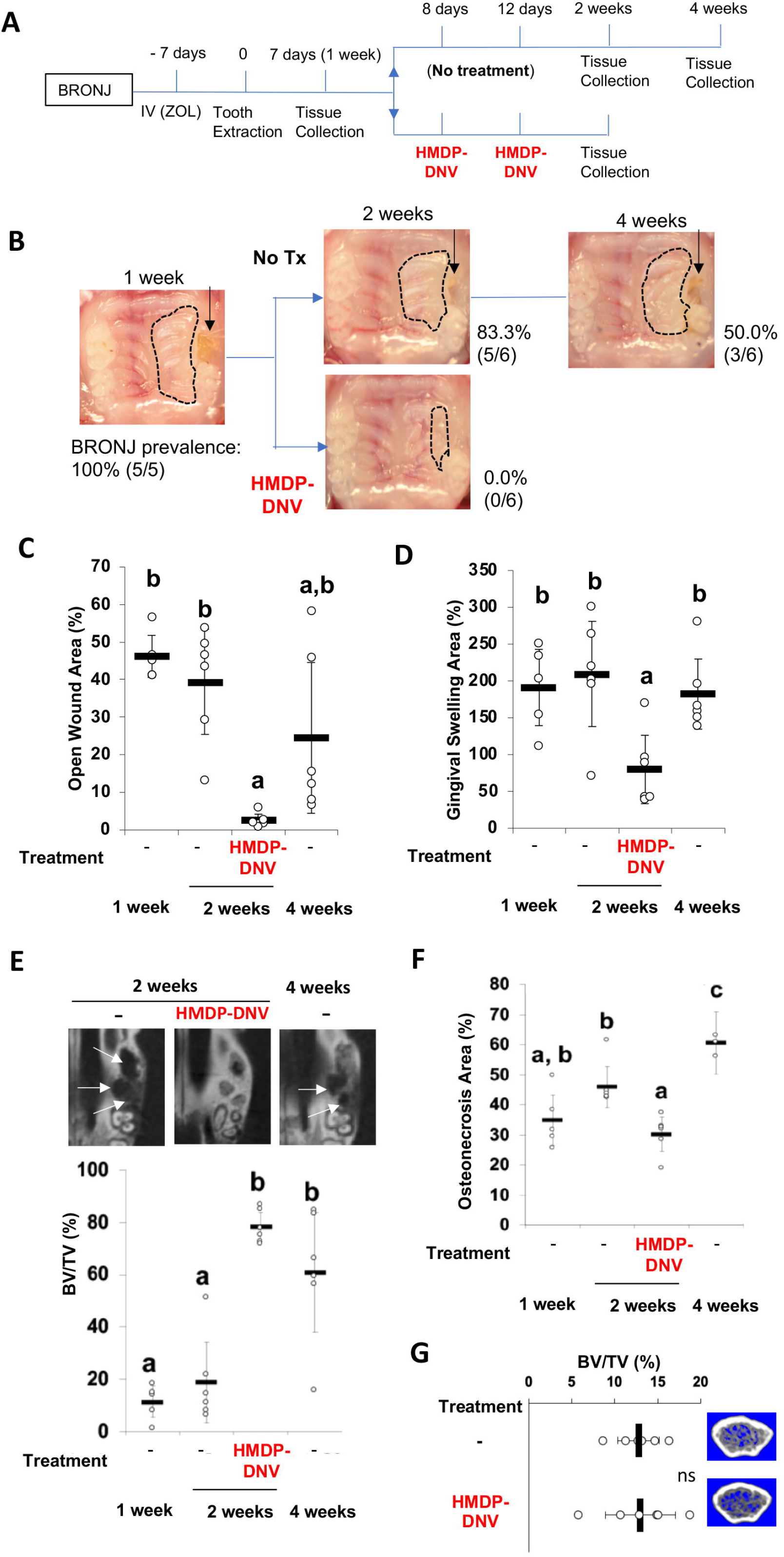
HMDP-DNV topical application to the established BRONJ lesion in mice accelerated the disease resolution. **(A)** The time course experimental protocol. After ZOL IV injection and tooth extraction, a BRONJ lesion developed, which was treated by two topical applications of HMDP-DNV in MQW (1.5 pmol/100 µM). **(B)** Intra-oral photographs depicted the unhealed tooth extraction wound (black arrows) with gingival swelling (dotted line) in significant percentage of animals in untreated mice group (‘No Tx’ group) from week 2 to 4. By contrast, all HMDP-DNV-treated mice exhibited a closed wound in week 2. **(C)** The wound opening of untreated mice remained 4 weeks after tooth extraction, while HMDP-DNV topical treatment minimized the wound opening 2 weeks after tooth extraction. **(D)** The gingival swelling area was also decreased by HMDP-DNV topical treatment. **(E)** Micro-CT analysis showed the delayed bone regeneration in the extraction sockets of untreated ZOL-injected mice. However, HMDP-DNV-treated ZOL-injected mice accelerated the extraction socket bone regeneration. **(F)** The histological osteonecrosis area progressively increased in the BRONJ lesion. HMDP-DNV topical treatment halted the osteonecrosis area increase. **(G)** The topical application of HMDP-DNV did not affect distant skeletal tissue in femurs.

Micro-CT imaging demonstrated the impaired bone regeneration in the extraction socket and extended osteolysis in BRONJ mice. By contrast, bone regeneration in the extraction socket was significantly improved by the HMDP-DNV treatment (Fig. 4E). The osteonecrosis area of maxillary alveolar bone at the tooth extraction site of the HMDP-DNV treated group was also significantly less than in the untreated BRONJ group at week 2 of extraction healing (Fig. 4F).

Micro-CT analysis of femur bones revealed increased trabecular bone volume and decreased inter-trabecular bone space in the ZOL-injected mice (Fig. 1B, 1C), as expected as the anti-resorptive effect of ZOL. Intra-oral topical application of HMDP-DNV did not alter the femur bone trabecular architecture, which exhibited typical phenotypes of the ZOL treatment (Fig. 4G). These results indicate that the treatment efficacy of HMDP-DNV was limited to the jawbone without modulating the antiresorptive effect of legacy ZOL in the other skeletal system.

### Oral barrier immune reaction examined by single cell RNA sequencing

Histological evaluations of mouse tooth extraction wound showed delayed bone healing and epithelial hyperplasia leading to the exposure of jawbone in ZOL-treated mice consistent with BRONJ lesion in humans. These histopathological events were attenuated in the HMDP-DNV treated mice (Fig. 5A). More notably, inflammatory cell infiltration to the gingival connective tissue was localized on the necrotic jawbone in the untreated mice. By contract, the inflammatory reaction was subsided, and the necrotic jawbone appeared to be actively removed by the normalized osteoclastic bone resorption (Fig. 5B).

**Fig. 5.**
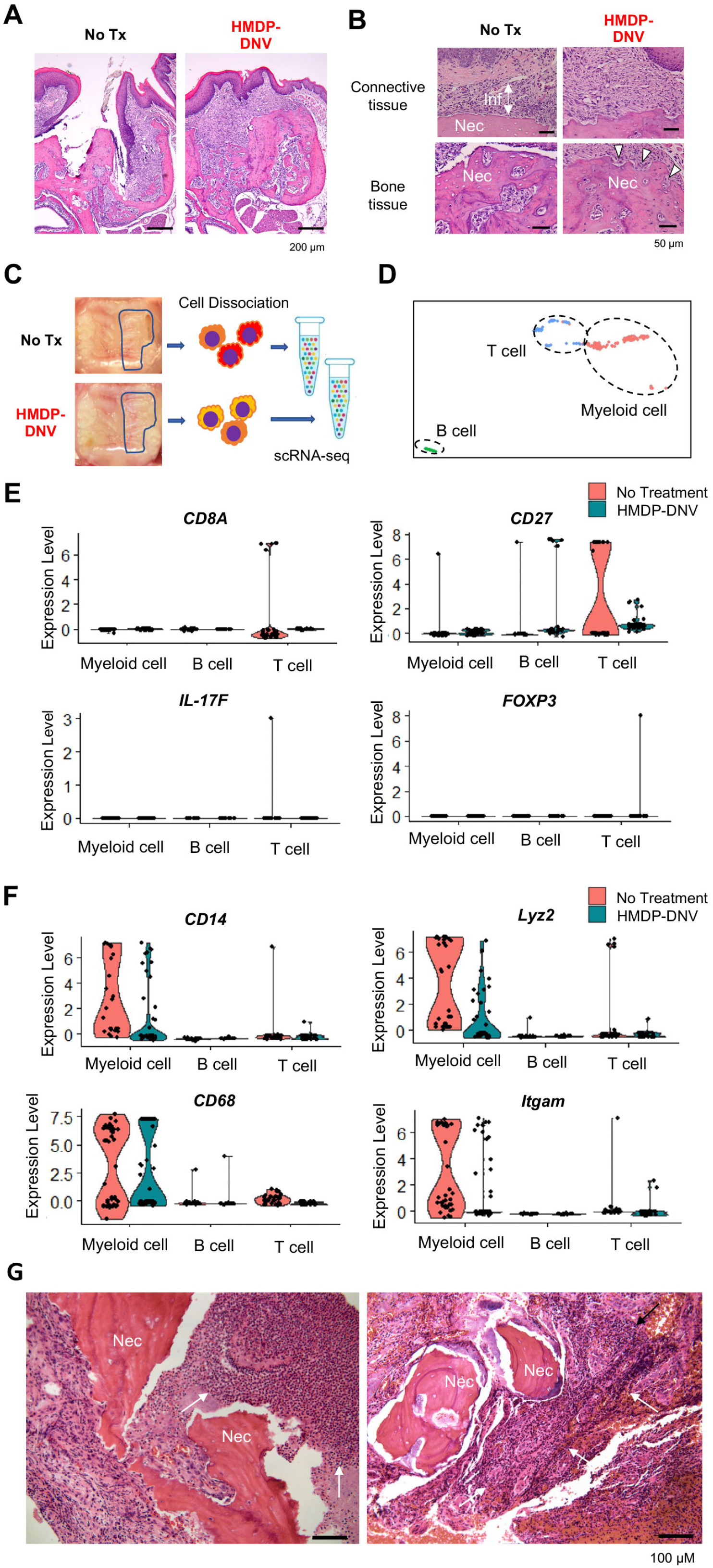
Single cell RNA sequencing of gingival cells of untreated and HMDP-DNV-treated ZOL-injected mice. **(A)** Histological evaluation depicted BRONJ lesion in the untreated mice (No Tx), which was significantly attenuated in HMDP-DNV-treated mice. **(B)** High magnification histology demonstrated the dense inflammatory cell infiltration (Inf) in the gingival connective tissue and osteonecrosis area (Nec) in untreated mice. The HMDP-DNV treatment attenuated the inflammatory cell infiltration and increased signs of osteoclastic bone resorption (white arrowheads). **(C)** Two weeks after tooth extraction, gingival tissue adjacent to the tooth extraction site was harvested for cell dissociation followed by single cell RNA-sequencing. **(D)** Using signature gene expression, myeloid cells, T cells and B cells were identified. **(E)** T cell-related gene expression indicated the presence of CD8A+ cytototoxic, CD27+ matured T cells derived from mouse BRONJ gingiva. IL-17F expression phenotype was decreased by HMDP-DNV treatment, which increased FOXP3 Treg phenotype. **(F)** Macrophage-related genes demonstrated an increase in M1 macrophages in untreated BRONJ gingiva, which was decreased by HMDP-DNV treatment. (**G**) Human BRONJ biopsy samples showing that necrotic bones (Nec) were associated with a large cluster of neutrophils (black arrows).

To determine the effect of HMDP-DNV treatment on the oral barrier immunity, we performed the single cell RNA sequencing of dissociated cells from gingival tissue (Fig. 5C). The gingival oral barrier cells were composed of T lymphocyte and B lymphocyte and myeloid cells (Fig. 5D). The fraction of T cells in the untreated BRONJ mice contained *CD8A*+ cytotoxic T cells and the upregulated expression of T cell costimulatory molecule CD27 (Fig. 5E), suggesting the presence of mature CD4+ and CD8+ T cell function (56). Furthermore, a small but distinct expression of *IL-17F* indicated the differentiation of highly pro-inflammatory Th17 cells (57) in the BRONJ lesion. The oral barrier cells from HMDP-DNV treated mice did not show these pro-inflammatory signatures. Instead, it was noted that the expression of *FOXP3* in their T cell fraction (Fig. 5E), suggesting the presence of regulatory T cells potentially mediating inflammation resolution (58) and wound healing (59).

Similarly, *CD14+Lyz2+* myeloid cells in the untreated BRONJ lesion showed the expression of *CD68* and *Itgam* (Fig. 5F), suggesting the involvement of macrophages in the innate immune system contributing to the pro-inflammatory condition (60). The attenuation or suppression of these pro-inflammatory functions was suggested in the HMDP-DNV treated oral barrier tissue. The histopathological evaluation of human BRONJ biopsy samples (Fig. 5G) as well as necropsy specimens from rodent BRONJ models (61, 62) reported the association with a large cluster of neutrophils.

### HMDP-DNV treatment induced myeloid-derived suppressor cell gene signature in the oral barrier tissue

The myeloid cell fraction of the scRNA-seq data were further divided into neutrophils and macrophages (Fig. 6A). It was striking that neutrophils in the HMDP-DNV treated oral barrier tissue lacked the expression of *TREM1* (Fig. 6B), which would trigger innate immune activation (63, 64). Furthermore, the expression of pro-inflammatory cytokines, *IL-1A*, *IL-1B* and *TNF* was nearly negated in neutrophils and macrophages by the HMDP-DNV treatment (Fig. 6C).

**Fig. 6.**
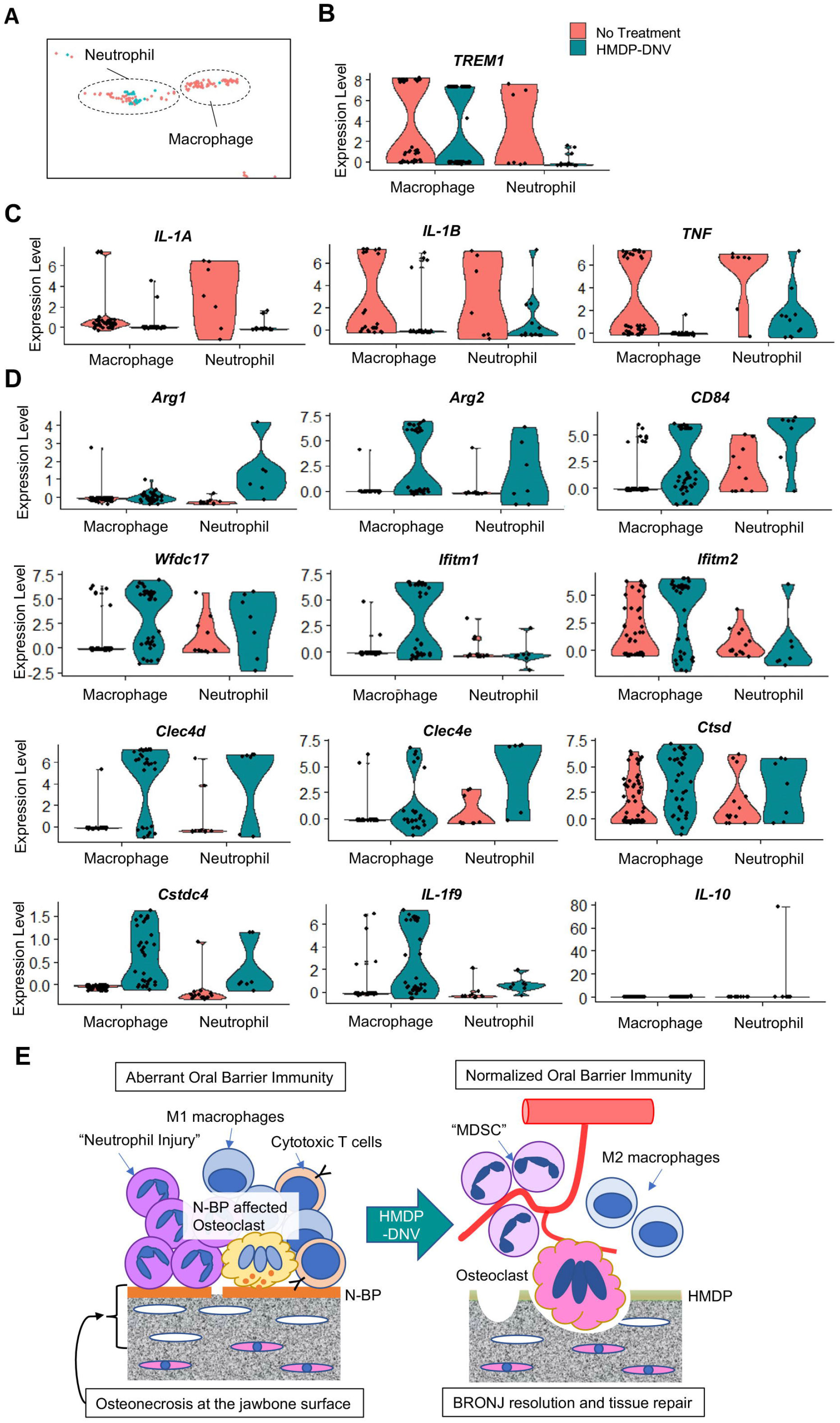
Characterization of myeloid immune cells. **(A)** The myeloid cell fraction of scRNA-seq was further divided into neutrophils and macrophages. **(B)** Myeloid immune cells identified by *TREM1* demonstrated a significant decrease of neutrophils by HMDP-DNV treatment. **(C)** Pro-inflammatory cytokines *IL-1A*, *IL-1B*, and *TNF* were highly expressed in macrophages and neutrophils from BRONJ gingiva. **(D)** Gingival macrophage and neutrophil myeloid cells after HMDP-DNV treatment expressed the multiple signature genes of the myeloid-derived suppressor cell (MDSC): *Arg1*, *Arg2*, *CD84*, *Wfdc17*, *Ifitm1*, *Ifitm2*, *Clec4d*, *Clec4e*, *Ctsd*, *Cstdc4* (from (69)) or M2 macrophage phenotypes *Arg1*, *Arg2*: as well as anti-inflammatory cytokines *IL-1f9* and *IL-10*. **(E)** A hypothetical model of BRONJ.

It was found that neutrophils as well as macrophages from HMDP-DNV treated oral barrier tissue expressed *Arg1* and *Arg2* (Fig. 6D), suggesting the stimulation of an anti-inflammatory reaction (65) and tissue regeneration/remodeling (66). *Arg*+ myeloid cells have also been known as myeloid-derived suppressor cells (MDSC) that negatively regulate the pro-inflammatory reaction and induce vascular development to support tissue regeneration and wound healing (67, 68). MDSC are diverse population of monocytic and granulocytic myeloid cells that are recently shown to express a common set of signature genes (69). We analyzed the scRNA seq data for the gene signature of MDSC. Macrophage and neutrophil myeloid cells isolated from the HMDP-DNV treated oral barrier tissues expressed all of the evaluated MDSC signature genes, including anti-inflammatory cytokines (Fig. 6D).

Taken together, the selective removal of ZOL by topical HMDP-DNV treatment from the jawbone prevented the progressive increase of osteonecrosis, normalized osteoclastic activity, and attenuated acute and chronic inflammation, ultimately leading to healing and resolution of the BRONJ lesion (Fig. 6E).

## DISCUSSION

The pathological mechanism of BRONJ has not been well established. This side effect of N-BPs selectively affects jawbone in the oral cavity. The unique features of the jawbone include its proximity to the oral immune barrier and frequent osteoclastogenesis caused by dentoalveolar infection, inflammation, and wounding (40, 70). We have hypothesized that the presence of N-BP on the jawbone is a critical causal factor, which interfaces the unique oral environment.

To examine our hypothesis, we developed a method to remove legacy N-BP only from the jawbone, particularly from the alveolar bone surface interfacing the oral barrier tissue. Unlike the common consensus, we found that N-BP chemisorption to HAp was not permanent, but rather a dynamic molecular equilibrium on the bone surface (43, 44). Thus, pre-adsorbed N-BPs can be displaced from HAp by exposure to a second N-BP/BP in the aqueous buffer. HMDP is a primary chemical component of Tc-99m-HMDP, a widely used diagnostic agent in Single Photon Emission Computed Tomography (SPECT) imaging. It is well established for its safety, and the selectivity of HMDP for SPECT imaging of abnormal bone metabolic sites (71) has been demonstrated. A recent clinical report indicated that Tc-99m-HMDP also binds efficiently to MRONJ lesions in humans (72). In view of these facts, HMDP was selected for this study, which might function as a benign agent for local removal of pre-adsorbed ZOL and normalizing osteoclastic activity (Fig. 1).

We previously reported that intra-oral injection of HMDP prevented the development of BRONJ in ZOL-injected mice (44). However, a single injection resulted in an uneven distribution of HMDP on the jawbone and a discrepancy in the effect at different locations (Fig. S4). To generate a more consistent effect, we developed the protocol for a new topical formulation of HMDP encapsulated in DNVs (HMDP-DNV) (Fig. 2). This new topical formulation also allowed the targeted and more consistent application of HMDP to the jawbone, while not interfering with other skeletal sites, making it possible to retain the systemic effect of legacy N-BP in the appendicular and vertebral bones. This new mouse model was used to elucidate the pathological mechanism of BRONJ.

Cochran review (CD012432, 2017) reported no standardized scale for the measurement of BRONJ. However, based on clinical examination, the extent of BRONJ was measured for the absolute area or percentage of initial area of wound opening. Other measures include the time taken for mucosa to completely cover necrotic tissue and exposed bone (“cure period”) (73).

This clinical oral wound observation was used to characterize the BRONJ resolution. A multicenter case registry study reported either resolution (35%) or improvement (10%) in 207 evaluable BRONJ patients during the study within the study period of 2 years with conservative treatments such as irrigation and antibiotic medications (74). However, 37% of patients did not respond to the conventional treatment and exhibited either progression or stable condition. A separate case series of 25 patients described that the natural resolution of BRONJ required necrotic bone sequestration and debridement leading to primary closure of the exposed bone (75). Cochran review (73) also reported the use of radiographic examination as the sign of BRONJ resolution.

A striking observation in this new mouse model was that HMDP-DNV therapy promptly assisted and accelerated wound healing and tissue remodeling of the mouse BRONJ lesion while preventing lesion enlargement (Fig. 3 and 4). Intra-oral administration of this therapy demonstrated wound closure and radiographic extraction socket bone healing with micro-CT in mice receiving the twice repeated HMDP-DNV topical application and demonstrated a rapid BRONJ resolution. The alveolar bone surface of BRONJ mice showed shallow bone resorption lacunae, likely due to the premature termination of osteoclastic activity (Fig. 3F, 5B). Topical HMDP-DNV treatment of the BRONJ lesion appeared to normalize osteoclastic bone resorption as evidenced by deep lacunae on the necrotic alveolar bone (Fig. 5B). The osteonecrosis area of the HMDP-DNV-treated BRONJ mice at week 2 did not increase from the week 1 osteonecrosis area before the treatment (Fig. 4F). The treatment effect of HMDP-DNV was not to revitalize the necrotic bone but to remove the affected bone by normalizing the osteoclastic activity and halting further necrosis.

Another observation was the resolution of a gingival inflammatory reaction. HMDP-DNV-induced removal of ZOL from the jawbone appeared to result in the reduction of gingival swelling (Fig. 4D) and inflammatory cell infiltration (Fig. 5A and 5B). To determine the mechanism of inflammation resolution, we developed a method of oral barrier cell dissociation suitable for single cell RNA sequencing. The single cell RNA sequencing analysis of gingival oral barrier immune cells clearly revealed the expression of pro-inflammatory signature genes in T cells and myeloid cells derived from BRONJ lesion. The HMDP-DNV treatment converted from this pro-inflammatory to anti-inflammatory phenotype (Fig. 5). A clinical biopsy report suggested that the necrotic bone was associated with bacterial aggregates, and severe inflammation (76) predominantly of neutrophils (Fig. 5G). In the present study, mice were not treated with antibiotics. Therefore, the resolution of pro-inflammatory reaction by HMDP-DNV treatment was not due to microbial suppression.

ZOL treatment was shown to decrease MDSC in mammary tumor-bearing female mice (77) and to decrease tumor-associated macrophages (TAM) in mesothelioma-bearing female mice (78). MDSC and TAM are important mediators of tumor-induced immunosuppression and the inhibition of MDSC accumulation with N-BP improves the host anti-tumor response in breast cancer (77) and pancreatic adenocarcinoma (79). MDSC has been highlighted in the tumor environment as pathologically activated neutrophils and monocytes with a detrimental role of enhancing tumor growth and metastasis (80). The present study demonstrated the clear gene signature of MDSC in the gingival oral barrier tissue after HMDP-DNV-derived removal of ZOL from the jawbone (Fig. 6). Because of the transient presence of MDSC during wound healing protects from over-reactive immune responses (81), MDSC in HMDP-DNV-treated mice may play an important role in the resolution of chronic inflammation leading to tissue repair promotion and the reestablishment of homeostasis.

It is unclear how jawbone N-BP affects the oral barrier chronic inflammation. The hematopoietic stem cell (HSC) niche is maintained by the bone marrow microenvironment. The relationship between HSC and mesenchymal stem cell (MSC) has been extensively investigated (82). However, N-BP treatment was strongly associated with delayed expansion of HSC, suggesting that normal osteoclast function was critical in the HSC niche (83). Osteoclasts are differentiated from myeloid immune precursor cells specialized for bone resorption. Recently, osteoclasts have been found to maintain the ability to secrete a set of anti-inflammatory cytokines (84, 85). N-BPs such as ZOL altered osteoclasts to increase the secretion of pro-inflammatory cytokines (85). Furthermore, osteoclasts are found to be an efficient feeder cells supporting natural killer cells in an *ex vivo* system (86). A cluster of osteoclasts induced by dentoaveolar infection and surgery may play a critical role in supporting and regulating the oral barrier immunity (87). Denosumab has been used in the same indications as N-BP through a different pharmacological mechanism; but denosumab-related ONJ (DRONJ) demonstrated similar clinical and radiographic symptoms (88). We postulate that the lack of viable osteoclasts as a local immune coordinator may cause the dysregulation of oral barrier immunity leading to the development of BRONJ and DRONJ.

In conclusion, the present studies of topical oral administration of HMDP-DNV demonstrated targeted removal of ZOL from the jawbone and provide a new mouse model to elucidate the mechanism of BRONJ. We underline that the local intra-oral application of HMDP-DNV did not affect the distant skeletal system and thus the pathological mechanism of BRONJ is likely localized within the oral tissue. The jawbone N-BP appears to adversely reduce oral osteoclasts, which may result in the dysregulation of the oral barrier immunity causing BRONJ. Finally, the work reported here establishes the basis for the development of this novel treatment as an effective prophylactic and therapeutic method for ZOL-induced BRONJ, and possibly for BRONJ associated with other N-BPs.

## MATERIALS AND METHODS

### Chemical reagents

Zoledronate (ZOL) was acquired from UCLA Medical Center Pharmacy (Reclast^®^, Novartis, Basel, Switzerland). Fluorescent-tagged ZOLs (FAM-ZOL and AF647-ZOL) were obtained from BioVinc LLC (Pasadena, CA). HDMP (oxidronic acid; hydroxymethylene-1,1-bis(phosphonic acid) was acquired from Aroz Technology (Cincinnati, US; Cat # BP-1026) as the disodium salt and characterized by ^1^H NMR (D_2_O, 600 MHz): δ3.78 (t, J = 16 Hz), ^31^P NMR (D_2_O, 243 MHz): δ 14.90 (s) and by elemental analysis: calculated for CH_4_O_7_P_2_Na_2_, 4.98% C, 1.92% H; found 4.97% C, 1.91% H (>99% purity). DOTAP (1,2-dioleoyloxy-3-(trimethylammonium)propane-sulfate), DPPC (diphosphatidylcholine), CH (cholesterol) and the nonionic surfactant Span 80 were acquired from Sigma-Aldrich (St. Louis, MO).

### The effect of ZOL and HMDP in mouse femur bone structure *in vivo*

Female C57BL/6J mice (n=5 per group) were anesthetized by isoflurane inhalation and 100 µl of ZOL (40 nmol), HMDP (40 nmol) or saline vehicle solution was injected to retro-orbital venous plexus (36, 61). Three weeks after the injection, mice were euthanized and femur bones were harvested for micro-CT imaging (µCT40, Scanco Medical AG, Southeastern, PA) following the standard procedure. Bone parameters were determined using the proprietary analysis program.

### Competitive displacement removal of ZOL by HMDP *in vitro*

Cell culture wells coated with carbonate apatite (Bone resorption assay plate 24, Cosmo Bio Co. Ltd, Tokyo, Japan) were incubated with fluorescent-tagged ZOL (10 µM FAM-ZOL,) overnight at 37°C, 2% CO_2_, followed by 3 washes with Milli-Q treated pure water (MQW) for 10 min each. The FAM-ZOL coated wells were then incubated with 10 µM HMDP in MQW (n=6) or MQW (N=3) for 2 hr at 37°C, 2% CO_2_ followed by 3 washes with MQW. One group of HMDP treated wells (n=3) were treated by the second application of 10 µM HMDP. Other wells were treated by MQW. After washes, the FAM fluorescent signal of each well was evaluated (IVIS Lumina II, PerkinElmer, Waltham, MA): excitation 465 nm; and emission filter: GFP (48). The region of interest was set to the well size and the fluorescent signal was measured. Separately, all wash solutions were subjected to fluorescent signal evaluation.

### Osteoclast resorption pit formation assay *in vitro*

Cell culture wells coated with carbonate apatite were incubated with ZOL (10 µM in MQW) for overnight at 37°C, 2% CO_2_ followed by extensive washes. ZOL-preincubated wells were treated with 10 µM HMDP once (n=3) or twice (n=3) as described above. Control wells were treated with MQW (n=3). After the final wash, culture medium (MEM containing 10% fetal bovine serum and 1% antibiotics mix) was added to all wells. Then, RAW274.1 cells (2.5×10^4^ cells per well) were inoculated to each well in culture medium supplemented by mouse recombinant receptor activator of nuclear kappa-B ligand (RANKL; Sigma-Aldrich) (100 ng/ml) and incubated at 37°C, 2% CO_2_. The culture medium was changed after 3 days. The resorption pits were photographed after 6 days of incubation after the cells were removed with 0.25% Tripsin. The total area of resorption pit was measured using a Java-based image processing program (ImageJ, NIH, Bethesda, MD).

### Synthesis of HMDP-DNV and AF647-ZOL-DNV

Fabrication of the DNV formulation containing BP compounds followed the published method (47). Briefly, cationic DOTAP lipid with DPPC and CH were dissolved in a 10 mM chloroform solution at a 5:3:2 volume ratio and the mixture was allowed to dryness. The dried lipid mixture was resuspended in isopropyl alcohol (final concentration 10 mM). To this solution, Span80 (15%v/v) was added. In the case of non-deformable DNV (nDNV), Span80 was not added. The lipid solution was filtered (0.2 µm) before injection in the reactor.

The aqueous solution was comprised of deionized water and the BP compound (12.9 µM) and filtered (0.2 µm). The organic and aqueous solutions were used for DNV synthesis using microfluidics reactor (Syrris, Royston, UK) with a 26 µL reactor chip at 1000 µL/min organic stream and 5000 µL/min aqueous stream, followed by dialysis to remove the non-encapsulated BP compound and free lipids and then freeze-dry processes. Physical characteristics such as the size and zeta potential were determined by a Malvern Zetasizer (Nano-ZS: Malvern, Worcestershire, UK) following the manufacturer’s protocol.

BP encapsulation efficiency (EE) was calculated as percent of AF647-ZOL amount in the DNV relative to the initial AF647-ZOL amount used in the synthesis. An aliquot of the lyophilized AF647-ZOL-DNV or AF647-ZOL-nDNV was lysed by adding acetonitrile. Next, samples were shaken for half an hour, then 350 μ compounds and samples were centrifuged for ten minutes at 22000xg. The supernatant was collected for LC. A standard curve was created using the original 12.9 µM stock solution of each of the compounds using LC for AF647-ZOL-DNV. Using the equation for the line of best fit for the signals received, we inputted the average signal given by the samples post-dialysis to obtain their concentration. Using the theoretical concentration of the samples after synthesis, we were able to calculate the encapsulation efficiency of the liposomes as 23%, which was verified (+/- 5%) by direct analysis of the unmodified HMDP-DNV formulation by a ^31^P NMR method (Supplemental file). It should be noted that the amount/concentration of all BP-DNV or BP-nDNV discussed in the following *in vitro* and *in vivo* studies were all calculated based on the encapsulated BPs in the DNV/nDNV formulation.

### LCMS analysis method for the encapsulation efficiency measurement of HMDP-DNV: Sample preparation

Five hundred (500) µL of ice-cold LCMS grade ACN was added to vial containing lyophilized HMDP-DNVs. The mixture was vortexed for 3 mins and sonicated for 20 mins to break the DNVs. This sequence was repeated once. Next, 2 µL LCMS grade water was added to each vial and the mixture was vortexed for 10 secs and centrifuged at 10,000 rpm for 30 mins.

Following the centrifugation, 1.5 µL of the supernatant was collected and separated from the pellet. A 5 mg/µL HMDP stock solution was prepared from the supernatant using maximum theoretical encapsulation value of 100 mg/vial. From the stock, 200 µL, 100 µL, and 40 µL corresponding to 1000 ng, 500 ng, and 200 ng HMDP respectively, were added to 1-dram glass vials. To each vial was added 200 µL of 500 ng/µL etidronate disodium solution as internal standard (IS). The samples were lyophilized. 500 µL trimethyl orthoacetate (TMOA) and 150 µL acetic acid was added to the lyophilized samples, and the mixture was heated at 100°c for 1h for derivatization. The solvents were evaporated under a stream of air. The residue was dissolved in 1 µL LCMS water. The samples were filtered, transferred to 1 µL HPLC vials, and the HMDP content was analyzed using LCMS. For generating standard curve, standard solutions were prepared by adding known concentrations of HMDP to 1-dram glass vials along with 200 µL of 500 ng/µL etidronate disodium as IS. The solutions were lyophilized, derivatized, and processed as mentioned above.

The LCMS system consisted of Waters Aquity H-class UPLC coupled with Waters Xevo G2-XS QTOF equipped with Masslynx 4.2 software (Waters, Massachusetts, USA).

Chromatography was performed on Syncronis ™ C18 Column (2.1 x 50 mM, particle size 1.7 mM, Thermo Scientific) using water (mobile phase A) and ACN (mobile phase B) at 30°C and 0.4 µL/min. The chromatography gradient was 0 - 0.3 mins (0% B), 0.3 - 0.8 mins (0 - 100% B), 0.8 - 1.6 mins (100% B), 1.6 - 2 mins (100 - 0% B), and 2 - 3.5 mins (0% B). The QTOF was operated in positive ion mode monitoring *m/z* transitions for methylated derivates of HMDP (313 > 161) and etidronate (327 > 267). Measurements of at least three different concentrations with 3x – 5x replicating tests at each concentration were performed, and the final EE value is calculated as the average value of these tests.

### Estimation of HMDP content in final DNV preparation by 31P NMR

To varify the HMDP content of HMDP-DNV, we conducted an independent analysis based on 31P NMR. Using the LCMS-based approach, the encapsulation efficiency (EE) was calculated as percent of HMDP amount in the DNV relative to the initial HMDP amount used in the synthesis. By this method, the final EE value for HMDP-DNV averaged to 23% which corresponds to a concentration of 3.0 µM for HMDP encapsulated by the liposome. Unlike the LCMS-based approach, ^31^P NMR analysis does not require chemical derivatization of the HMDP. Thus, an aliquot of the HMDP-DNV formulation was used directly as the sample. The average integration of the ^31^P NMR signal for an accurately weighed sample of HMDP in D_2_O was determined relative to the signal of an external reference of known concentration (analytically pure methylenebisphosphonic acid, disodium salt). The same system was then used to determine the concentration of HMDP in the HMDP-DNV formulation, in D_2_O. The value obtained by the latter method was 2.7 µM, which agreed within error with the value of 3.0 µM from the LCMS analysis, giving an encapsulation efficiency of 23% for a starting HMDP concentration of 12.9 µM in the HMDP-DNV synthesis.

### Topical drug application to the mouse palatal mucosa and AF647-ZOL-DNV delivery to the jawbone

We designed a topical application protocol for delivery of BP-DNV to the maxillary jawbone through palatal mucosa. Mouse oral appliances were fabricated using clear orthodontic dental resin to fit to the entire mouse palatal mucosa surface between the molar teeth. Mice were anesthetized by isoflurane inhalation and placed on a surgical bed to open the mandible. Three (3) µl of AF647-ZOL-DNV or AF647-ZOL-nDNV reconstituted in MQW or 20% polyethylene glycol solution was pipetted to the palatal mucosa and covered by an oral appliance. The mandible was closed with a bite block and the animal was placed in the anesthesia chamber. Mice were kept anesthetized for 1 hour and the oral appliance was then removed. After 24-48 h, mouse maxillae were harvested and the AF647 fluorescent signal was measured by an imaging reader (LAS-3000, Fujifilm Corp, Tokyo, Japan).

### Induction and characterization of BRONJ lesion in the mouse model

Female C57Bl6/J mice (8 to 10 weeks of age) received a bolus intravenous injection of ZOL (500 µg/Kg in saline solution) or vehicle saline solution from the retro-orbital venous plexus. One (1) week later, the maxillary left first molar was extracted under general anesthesia by isoflurane inhalation (36, 61). After tooth extraction, all mice were fed gel food (DietGel Recovery, Clear H_2_O, Portland, ME) for 1 week and then switched to regular mouse pellet chaw. After switching to the regular pellet chaw, all mice were briefly anesthetized once a week and food and debris impaction was removed. Mice were euthanized at 1 week, 2 weeks or 4 weeks after tooth extraction and femur bones and the maxillary tissues including the extraction wound were harvested after being photographed. The buffered formalin-fixed maxillary tissues and femur bones underwent Micro-CT imaging. The maxillary tissues were demineralized by 1 M EDTA in 4°C and prepared for paraffin-embedded histological sections stained with hematoxylin and eosin. Digitized histological images were examined and the area of osteonecrosis was measured using the ImageJ program. The mouse oral lesion, Micro-CT radiography and histopathology were compared to clinical information from human BRONJ patients.

### Topical application of HMDP-DNV to the mouse BRONJ model prior to tooth extraction

ZOL-injected mice were treated with topical application of Empty-DNV, HMDP in MQW (1.5 pmol/100 µM) or HMDP-DNV in MQW (1.5 pmol/100 µM) to the palatal oral mucosa. The topical formulation was applied 1 time (Figure S3A) or 2 times (Figure 3H) in a week and then the maxillary left first molar was extracted. Two (2) weeks after tooth extraction, mouse maxillae and femur bones were harvested. The development of BRONJ lesion was examined as described above.

### Topical application of HMDP-DNV to the established mouse BRONJ lesion

ZOL-injected mice underwent a maxillary left first molar extraction. Delayed healing and BRONJ-like jawbone exposure were confirmed 1 week after tooth extraction. HMDP-DNV in 3 µl MQW (1.5 pmol/100 µM) was topically applied to the BRONJ lesion and surrounding gingival tissue. This treatment was applied 2 times total in a week. The maxillary tissues and femur bones were harvested and the BRONJ lesion was characterized as described above.

### Single cell RNA sequencing of gingival oral barrier immune cells

After the HMDP-DNV treatment of the BRONJ mice, the gingival tissue from the tooth extraction side of the palate was harvested (n=4). The gingival tissue of untreated BRONJ mice was also harvested (n=4). Gingival tissues were cut into 1-mm pieces and placed in digestion buffer containing 1 mg/ml collagenase II (Life Technologies, Grand Island, NY), 10 units/ml DNase I (Sigma-Aldrich) and 1% bovine serum albumin (BSA; Sigma-Aldrich) in Dulbecco’s modified Eagle’s medium (DMEM; Life Technologies) for 20 min at 37°C on a 150 rpm shaker. The tissues were passed through a 70µm cell strainer. The collected cells were pelleted at 1,500 rpm for 10 min at 4°C and resuspended in phosphate buffered saline (PBS; Life Technologies) that was supplemented with 0.04% BSA (Cell suspension A).

The remaining tissues were further incubated in 0.25% trypsin (Life Technologies) and 10 units/ml DNase I for 30 min at 37 LJ on a 150 rpm shaker. The combined trypsin-released cells and collagenase II-released cells were counted and the equal number of cells (2,000∼3,000) from each animal were combined in the group for single cell RNA sequencing (10X Genomics, San Francisco, CA). The Cell Ranger output of single cell RNA sequencing data were analyzed using R-program (Seurat, https://satijalab.org/seurat/).

### Statistical analysis

The mean and standard deviation were used to describe the data. The Turkey test was used to analyze multiple samples and statistical significance was considered to be achieved if p<0.05.

### Study approval

All animal experiments were performed at UCLA. All the protocols for animal experiments were approved by the UCLA Animal Research Committee (ARC# 1997-136) and followed the Public Health Service Policy for the Humane Care and Use of Laboratory Animals and the UCLA Animal Care and Use Training Manual guidelines. The C57Bl/6J mice (Jackson Laboratory) were used in this study. Animals consumed gel or regular food for rodents and water *ad libitum* and were maintained in regular housing conditions with a 12-hour-light/dark cycles at the Division of Laboratory Animal Medicine at UCLA.

Clinical demonstration of human BRONJ was obtained from patients of UCLA School of Dentistry clinic with the general consent for educational use. The information was not part of investigator-initiated research.

## Data Availability

All data produced in the present work are contained in the manuscript.

## Author contributions

Conceptualization: IN, AH, CEM, SS, FHE

*In vitro* and *in vivo* experiments and data collection: HO, TK, AH, IN

Funding acquisition: IN, FHE, CEM, HO, TK

Single cell RNA sequencing: TK, IN

Synthesis and characterization of DNV formulation: JJC, VJ, SS, PC, NL

Summary of MRONJ prevalence data: SC

Patient case reports: ECS, Y-LL

Data analysis: IN, AH, HO, TK, SS, FHE

Writing – original draft: IN, CEM with contributions from AH; revision and final version: IN, CEM, SS, FHE

## Acknowledgments

We thank Dr. Hodaka Sasaki, UCLA School of Dentistry and Tokyo Dental College for designing the mouse oral appliance, Dr. Jimmy Hu of UCLA School of Dentistry for his guidance in single cell RNA sequence analysis, the UCLA Translational Pathology Core Laboratory for histological specimen preparation, the UCLA Technology Center for Genomics & Bioinformatics for 10XGenomics single cell RNA sequencing, and Inah Kang, USC for skilled assistance in preparing the manuscript.

## Supplementary Material

Fig. S1. Bisphosphonate-related osteonecrosis of the jaw (BRONJ) cases.

Fig. S2. Mouse BRONJ model characterized by Micro-CT.

Fig. S3. Single intra-oral topical application of HMDP-DNV to ZOL-treated mice prior to tooth extraction did not prevent development of a BRONJ lesion.

Fig. S4. HMDP intra-oral injection and HMDP-DNV topical application.

